# Why were Twitter Users Obsessed with Vitamin D during the first year of the pandemic?

**DOI:** 10.1101/2022.04.13.22273830

**Authors:** Alexandra Mavroeidi, Ryan Innes, Esperanza Miyake, Diane Pennington

**Author notes:** <u>Corresponding author:</u> Dr Alexandra Mavroeidi, Senior Lecturer in Physical Activity for Health, School of Psychological Sciences & Health, University of Strathclyde, Level 5, Graham Hills Building (room 538), Glasgow G1 1QE.

## Abstract

The aim of this study was to explore how the relationship between vitamin D and COVID-19 has been represented on the social media site Twitter. NCapture was used to collect textual Tweets on a weekly basis for three months during the pandemic. In total, 21,140 Tweets containing the keywords “vitamin D” and “COVID” were collected and imported to NVivo12. An inductive thematic analysis was carried out on the Tweets collected on the first (12/2/2021) and last week (21/5/2021) of the recording period to identify themes and subthemes. Quality control of the coding was conducted on a sample of the dataset (20%). Data were also compared to the “ground truth” to explore the accuracy of media outputs. The four main themes identified were “association of vitamin D with COVID-19”, “politically informed views”, “vitamin D deficiency” and “vitamin D sources”. When compared to the ground truth, the majority of information relating to the key findings was ‘incorrect’ for all of the findings. This study contributes to the area of research by highlighting the extent of the issue social media sites face with health-related misinformation. In the context of COVID-19, it is important that sites such as Twitter improve their existing misinformation policies, as misinformation can be detrimental in disease prevention.

## Introduction

The COVID-19 pandemic caused by the severe acute respiratory syndrome coronavirus 2 (SARS-CoV-2) is a subject of global concern (Misra et al., 2020). There has been an unprecedented response across several sectors to develop targeted therapeutics to slow the rate of infection (Carter et al., 2020). One modifiable lifestyle intervention that has received particular interest is vitamin D (Lanham-New et al., 2020).

The scientific debate basis for the potential relationship of vitamin D status with COVID-19 is based upon the association of low serum concentration of 25(OH)D with increased susceptibility to acute respiratory tract infections (Ilie et al., 2020). Several correlational studies have reported a relationship between vitamin D deficiency and poor COVID-19 outcomes (Ilie et al., 2020; Sulli et al., 2021; Baktash et al., 2020). However, a number of risk factors for poor COVID-19 outcomes are similar to those for vitamin D deficiency such as old age and darker skin (The World Health Organization, 2020; Dey & Sinha, 2020). Consequently, there are several other risk factors that must be considered before the true extent of the cause-and-effect relationship between vitamin D and COVID-19 can be extrapolated.

The potential impacts of social media on the COVID-19 pandemic are currently unknown, as it is the first pandemic to develop in the era of mass social media use. Social media differs to traditional media as it allows users to both consume and contribute to news in real time (Das & Ahmed, 2020). This participatory culture in news and social media could have both positive and negative consequences (Almgren & Olsson, 2016; Robinson & Wang, 2018). Factually correct information can be circulated – by official government sources e.g., Scottish Government (https://twitter.com/scotgov) - on social media faster than is possible with traditional media sources. Sharing accurate information at the earliest time possible such as local guidelines can help reduce the rate of infection (Mejia et al., 2020). However, social media is also synonymous with misinformation (Mancosu & Vegetti, 2020; Shu et al., 2020). The WHO stated that there was not only a pandemic, but also an “infodemic” which was defined as an outbreak of misinformation causing mass anxiety and uncertainty (Das & Ahmed, 2020). In the pandemic response, misinformation is dangerous, as information is a key factor in disease prevention (Mejia et al., 2020).

The aim of this study was to explore how the relationship between vitamin D and COVID-19 was presented on the social media site Twitter during the first year of the pandemic, and assess the level of misinformation presented in Twitter around this issue.

## Methods

Ethical approval was obtained through the Strathclyde University Computer and Information Sciences Ethics Committee on 1/2/2021 (ID: 1303).

### Data Collection

Textual data were collected from the social media site Twitter. NCapture (NVivo 12 plugin) was used to collect all Tweets in English containing the keywords “vitamin D” and “COVID” using the advanced search function on Twitter; no hashtags were used. NCapture does not have the ability to capture images, videos or emojis, so these were not included in the analysis. This was also the main reason why other platforms such as Instagram and TikTok were not included in this analysis, as they mainly contain image and video data. Facebook was also not included because its content is not considered publicly available and thus there are additional ethical considerations. Tweets were collected on a weekly basis for three months from 12/2/2021 until 21/5/2021- resulting in a total of 21,140 Tweets being collected. This time period was chosen as vaccination programmes were rolled out in most countries in the developed world, including the UK. As a consequence, alternative preventative remedies were a trending topic on Twitter. Collecting Tweets on a weekly basis made it possible to track the prevalence of Tweets containing the keywords (Figure 1).

**Figure 1.**
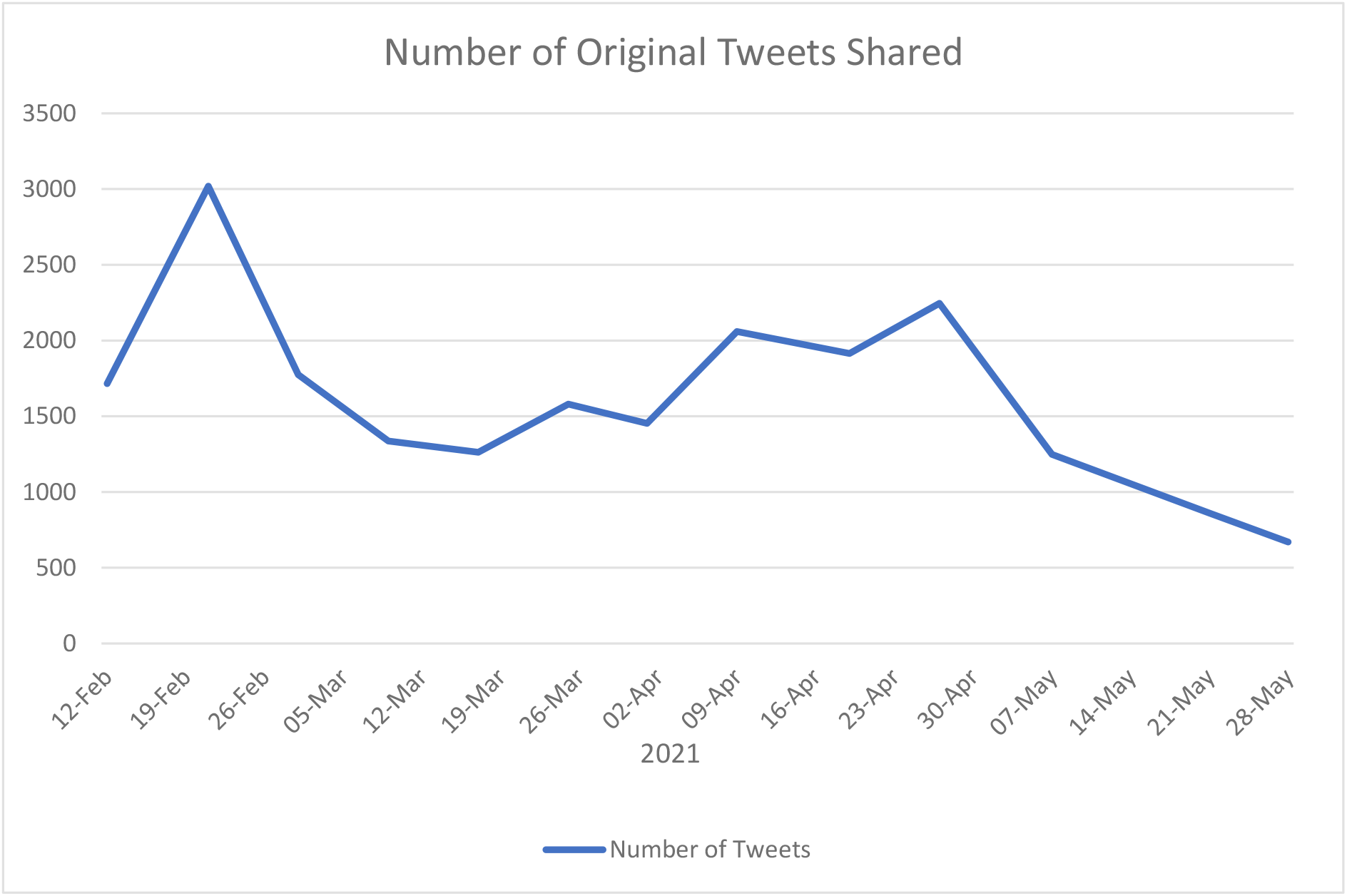
Number of Original Tweets Shared containing the keywords “Vitamin D” and “COVID”. Retrieved 9/4/21.

### Data Analysis

Inductive thematic analysis was used to investigate how the relationship between vitamin D and COVID-19 has been represented on Twitter. Thematic analysis is a qualitative technique that enables researchers to extract meanings and concepts from data sets that cannot be quantitively analysed, such as focus group transcripts (Javadi & Zarea, 2016). The technique is commonly utilized in health settings to analyse patient experiences (Hewis, 2015; Attard & Coulson, 2016; Lane et al., 2016). An inductive approach was used, meaning that the analysis was data driven (Braun & Clarke, 2006). Therefore, the themes identified are strongly linked to the data as the student researcher (RI) coded the data without trying to fit it into a pre-existing coding frame (Braun & Clarke, 2006). The student researcher (RI) followed the step-by-step guide outlined in (Braun & Clarke, 2006) to analyse the data. The six steps are as follows: familiarising yourself with your data, generating initial codes, searching for themes, reviewing themes, defining and naming themes, and producing the report.

The inductive thematic analysis was carried out in NVivo 12 - a software package commonly used for qualitative analysis (Edhlund & McDougall, 2019). Only Tweets collected on 12/2/2021 and 21/5/2021 were analysed due to time constraints (Appendix A & B). Quality control of the coding was conducted by another postgraduate research student with previous experience of thematic analysis (anonymous 1), who coded a sample of the dataset (20%). The student who carried out the quality control (anonymous 1) agreed with the provisional themes and sub themes identified by the student researcher (anonymous 2).

The codes identified from analysis of the Twitter data set collected on 12/2/2021 were also compared to the “ground truth” in order to explore the accuracy of media outputs. The NICE report titled “COVID-19 rapid guideline: vitamin D” was used as the ‘ground truth’ (National Institute for Health and Care Excellence, 2020). The four key findings identified from the NICE report were:

1. Do not offer a vitamin D supplement to people solely to treat COVID-19.
2. Do not offer a vitamin D supplement to people solely to prevent COVID-19.
3. Low vitamin D status was associated with more severe outcomes from COVID-19. However, it is not possible to confirm causality because many of the risk factors for severe COVID-19 outcomes are the same as the risk factors for low vitamin D status.
4. Adults, young people and children over 4 years should consider taking a daily supplement containing 10 micrograms of vitamin D between October and early March. Groups at high risk of vitamin D deficiency should also consider taking a daily supplement containing 10 micrograms of vitamin D year-round.

Data that did not relate to the key findings outlined above were excluded from this portion of the analysis. Appropriate data were compared to the key findings and coded in NVivo into either ‘correct’ or ‘incorrect’ nodes.

## Results

### Descriptive

The number of original Tweets shared containing the keywords “vitamin D” and “COVID” peaked in mid-February before steadily declining until mid-March, at which point, the number of Tweets generally trended upwards until late April. The number of Tweets then rapidly declined over the month of May. (Figure 1).

Analysis of the Tweets collected on 12/2/2021 and 21/5/2021 resulted in four main themes and twelve subthemes being identified (Table 1, Appendix A, B & C).

**Table 1.** Summary of themes and subthemes for Twitter Data.

| Theme | Subtheme |
| --- | --- |
| Association of Vitamin D with COVID-19 | COVID-19 Infection<br>COVID-19 Outcomes<br>COVID-19 Treatment |
| Politically Informed Views | Government Measures<br>Media<br>Conspiracy Theories |
| Vitamin D Deficiency | High Risk Groups<br>Impact of Vitamin D on Health<br>Prevalence of Vitamin D Deficiency |
| Vitamin D Sources | Food<br>Sunlight<br>Supplementation |

### Theme 1: Association of Vitamin D with COVID-19

#### Subtheme 1A: Covid-19 Infection

The vast majority of Tweets shared the incorrect belief that vitamin D was directly linked with COVID-19 infection.

> *“@redacted Have heard 2 different seasoned and previously reliable physicians state that they have never seen a Covid case in a person with adequate levels of vitamin D. So simple.” (Twitter User 1)*
>
> *“@redacted And yet the document makes no mention of supplementation especially vitamin -D. Doesn’t everyone at this point know about the importance of vitamin-d supplementation in covid prevention. Shouldn’t that be in there?” (Twitter User 2)*

The belief that scientific literature had ‘proven’ the relationship between vitamin D and COVID-19 infection was also common.

> *“@redacted There is by now so much literature that vitamine D deficiency exacerbates the chance to get infected by COVID-19 or a worse outcome, that there is no sane reason one can think of not to make combatting vitamin D deficiency an urgent priority this pandemic.” (Twitter User 3)*

#### Subtheme 1B: COVID-19 Outcomes

Users frequently cited results from published research that indicate a correlation between vitamin D status and COVID-19 outcomes.

> *“Heidelberg study shows Vitamin D deficiency correlated with increased incidence of ventilation & death for Covid patients: https://t.co/BeFZIB9tWE* https://t.co/c7DAR0MDgr*” (Twitter User 4)*
>
> *“I may not understand the math used here, but the results line up with other research which shows the association of better levels of vitamin d with better outcomes in Covid-19 infections. This is a tool that can improve overall immunity; we need to hear more about it*. https://t.co/5zgBtCvY7T*” (Twitter User 5)*

Tweeters shared their personal experiences with COVID-19, with many believing that vitamin D status played a role in their own illness.

> *“I hd covid - ws taking a number of suppliments prior 2 getting it. I think I got sick mostly due to lack of sleep due 2 the heat wave. Lack of sleep can compromise your immune system. But I hd zero lung infection just a fever - no coughing - notably I ws taking vitamin D #covid” (Twitter User 6)*
>
> *“Twitter/Facebook censoring medical information deserves a class action lawsuit. Vitamin D made my Covid-19 experience a stuffy head. VitaminD likely has the same effect on Cold/Flu. The censorship is malpractice by proxy. @redacted” (Twitter User 7)*

#### Subtheme 1C: COVID-19 Treatment

Users commonly recommended vitamin D as a COVID-19 treatment per se, rather than an aiding dietary supplement.

> *“@redacted Hi redacted can I suggest taking a look at niacin flush as well as vitamin d for treatment of covid 19 symptoms.” (Twitter User 8)*
>
> *“@redacted I had COVID in October, feeling great now, take vitamin D and stop in bed, you will likely feel better soon.” (Twitter User 9)*

However, there were also users that highlighted vitamin D was not a proven treatment for COVID-19.

> *“@redacted Agreed that Vitamin D is good for you in appropriate amounts, but as far Covid treatment is concerned, it and Ivermectin seem more like false promise wrapped in a thin layer of anecdote and rationalisation.” (Twitter User 10)*
>
> *“@redacted There is no evidence that vitamin D has anything to do with COVID. It certainly doesn’t seem to have any impact on treating it.” (Twitter User 11)*

### Theme 2: Politically Informed Views

#### Subtheme 2A: Government Measures

Lockdown measures and the restrictive effect that these might have had to people’s exposure to sunlight, and thus diminishing vitamin D status, were heavily criticised.

> *“*@redacted For morons out there that don’t get my joke, lock downs are moronic, cause more harm than good. While it’s all theories with COVID, will be interesting to see if low Vitamin D caused bad outcomes, because that would mean the lockdowns more adversly affected dark skinned people.” (Twitter User 12)
>
> *“Oh so they want to say now there is a link of low Vitamin D levels (lack of sunshine) raises the risk of covid/death YOU LOCKED EVERYONE AWAY FROM IT FUCKERS 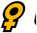 UK keeping research on link between vitamin D and Covid under review” (Twitter User 13)*

There was also a widespread belief that face masks were ineffective in reducing the risk of COVID-19 transmission.

> *“@redacted MASKS DONT WORK*. *Eat healthy, exercise, take vitamins, VITAMIN D especially (it’s proven to fit Covid)*. *I don’t wear a mask, and guess what I haven’t gotten Covid.” (Twitter User 14)*
>
> *“@redacted Begging for something that could damage or kill you. People who die of covid are vitamin D and zinc deficient. A mask doesn’t work and this vaccine could cause severe reactions to other minor illnesses. You were warned.” (Twitter User 15)*

#### Subtheme 2B: Media

Media originations were criticised for their apparent lack of coverage of the potential association of vitamin D with COVID-19.

> *“Has the MSM ever mentioned the impact of Vitamin D deficiency with COVID-19?” (Twitter User 16)*
>
> *“You can find over a dozen studies that show that vitamin D deficiency is a huge factor in Covid cases. And CB you can find multiple studies that show it is a great way to fortify your immune system. Hardly a mention in MSM try to find the studies about B masks. Good luck” (Twitter User 17)*

Trust in the media also appeared to be low.

> *“@redacted The MSM has their orders. We’re being culled and prepped for something evil. Especially our health care providers.” (Twitter User 18)*

#### Subtheme 2C: Conspiracy Theories

It was commonly theorized that information regarding the benefits of vitamin D in relation to COVID-19 was being suppressed for monetary purposes.

> *“@redacted According to a study from Spain injecting a high level of vitamin D into the bloodstream makes Covid hospital patients 50 times less likely to end up in ICU than those who took the placebo. But there’s NO MONEY TO BE MADE FROM VITAMIN D so won’t happen here in the UK.” (Twitter User 19)*
>
> *“@redacted Vitamin D is known to ward off respiratory viruses, including covid, but Big Pharma doesn’t want that to get around because an illness prevented (like a patient cured) is another customer lost.” (Twitter User 20)*

COVID-19 vaccines were also often described in a negative light (e.g. unsafe and useless) while vitamin D was presented as the miracle safe cure for COVID.

> “Instead of injecting people with an unknown, unsafe vaccine. Distribute vitamin D, C, zinc and HCQ. It’s healthy, affordable, safe & beats covid. Since they’ve lied, manipulated and used taxpayers money to fund this farce throw in a basket of fruits & veggies & a fat cheque.” (Twitter User 21)
>
> *“@redacted So why take a vaccine that’s useless? Better to take vitamin D #COVID19” (Twitter User 22)*

### Theme 3: Vitamin D Deficiency

#### Subtheme 3A: High Risk Groups

Three groups received particular attention: the elderly, BAME and obese individuals.

> *“Vitamin D supplementation to the older adult population in Germany has the cost-saving potential of preventing almost 30,000 cancer deaths per year. Again, irrespective of Covid (and indications are that D3 is a GREAT idea for that) all older people, D3!” (Twitter User 23)*
>
> *“@redacted There is an argument to be made that your skin color affects your outcome. It’s well known that the darker your skin, the less Vitamin D you get from sunlight.” (Twitter User 24)*
>
> *“Obesity is strongly correlated with Vitamin D deficiency. Vitamin D deficiency is a major factor in whether someone becomes seriously ill or dies from Covid. Vitamin D is fat soluble and gets stored in fat cells instead of circulating in your blood where it can do you some good.” (Twitter User 25)*

#### Subtheme 3B: Impact of Vitamin D on Health

Tweets frequently mentioned the important role vitamin D plays in the maintenance of general health.

> *“While it’s extremely important for the peer reviewed clinical trial evidence to be published before medical decisions are made using the information, there are other reasons why being deficient in vitamin D is unhealthy #COVID” (Twitter User 26)*

The positive impact on general health was linked to improved outcomes in COVID- 19 sufferers.

> *“@redacted Her point is losing weight, keeping fit, taking vitamin D etc will help in fighting off Covid, which is true but very rarely mentioned in the media.” (Twitter User 27)*

However, it was also pointed out that vitamin D status alone does not determine if an individual is healthy.

> *“@redacted Vitamin D important in context of overall Health. Overall Health significant in lessening severity of COVID. But overall health is more than Vitamin D, it is not a therapy for COVID or alternative to vaccines and will be unable to protect those who most need protection from COVID” (Twitter User 28)*

#### Subtheme 3C: Prevalence of Vitamin D Deficiency

The proportion of the population that are reportedly vitamin D deficient varied greatly in Tweets.

> *“@redacted I think nearly everyone is deficient of Vitamin D. What I would like to see in data is if they ever check levels of acidic vs alkaline balanced people that 1. contract covid and 2. how they recover but probably too holistic for them to do.” (Twitter User 29)*
>
> *“@redacted Absolutely, from the beginning, around March/April, I heard about vitamin d and zinc. Did you know deficiencies in zinc and vitamin d are relatively rare among the populace but the majority of people hospitalized with covid are deficient?” (Twitter User 30)*

### Theme 4: Vitamin D Sources

#### Subtheme 4A: Food

Food fortification programs were linked to COVID-19 outcomes.

> *“Higgins points to Finland, which fortifies food with vitamin D and has the lowest Covid death rate in Europe aside from Iceland” (Twitter User 31)*
>
> *“@redacted New Zealand also has a national vitamin D program and supplements their food with it. That vitamin has demonstrated that it lowers incidence, severity and fatalities in COVID.” (Twitter User 32)*

Foods’s rich in vitamin D were often listed with claims that they could prevent mortality in COVID-19 sufferers.

> *“@redacted But aren’t egg yolks a good source of vitamin D, which I’m pretty sure the Mail told me reduces Covid mortality?” (Twitter User 33)*
>
> *“VITAMIN D*
>
> *PEOPLE WHO DIED FROM COVID HAD LOW VITAMIN D*
>
> *GET OUT IN THE SUNSHINE, DRINK OJ, EAT SALMON, CHEESE, EGGS, SOY MILK, MUSHROOMS AND DAIRY” (Twitter User 34)*

#### Subtheme 4B: Sunlight

Several Twitter users believed that vitamin D obtained through sunlight exposure could ‘fight’ COVID-19.

> *“@redacted how about you stop with the masks and tell people to get out and soak up some sun. ultra violet light helps produce vitamin D and vitamin D enhances our immune system and helps fight covid” (Twitter User 35)*
>
> *“@redacted night walking reduces exposure to sunlight. This reduces vitamin d production which impinges upon the effectiveness of the immune system. During covid, people should walk in daylight to improve chances of fighting the disease and improving survival chances.” (Twitter User 36)*

There was also particular interest in the homeless population due to their increased exposure to sunlight.

> *“@redacted The only thing the homeless have going for them is plenty of sunshine and vitamin D. Maybe some “scientist” should pick up on that as they are virtually immune to Covid.” (Twitter User 37)*
>
> *“@redacted Am I reading this right? I think they’re saying why aren’t homeless people dying - well, they’ve probably got very good vitamin D levels, much like those farm workers who all had covid and didn’t know. The people who are dying are old people who don’t get out enough” (Twitter User 38)*

#### Subtheme 4C: Supplementation

There was a widespread belief that the government recommended dose of 10 micrograms was too low for both general health and any perceived COVID-19 benefits.

> *“@redacted RDA is typically grossly underestimated (I wonder why that it is?). 5,000iu D3 to maintain your existing level. 10,000iu to increase your D3 level.” (Twitter User 39)*
>
> *“As reported on the front page of every mainstream newspaper! Oh. Maybe not*
>
> *Habitual use of a decent level of Vitamin D (no, not 400iu that will do nothing for you) reduces COVID infection by 34%*
>
> *@redacted #vitaminD #COVID19 #KBF” (Twitter User 40)*

Twitter users also commonly stated that they were personally taking a vitamin D supplement that was much higher than the 10 micrograms recommended.

> *“Take vitamin D to hugely increase your chances of not getting covid and of surviving if you do get it. The dose should be at least 3,000 iu (international units). I take 10,000 iu. Doesn’t Witty know this?” (Twitter User 41)*
>
> *“@redacted I was fit and healthy and took many vitamins including 20000 iu vitamin D and it’s partner in crime K2 daily but I got Covid bad 14 days in bed and then 3 further weeks to recover not fully recovered yet” (Twitter User 42)*

Some users highlighted that even if vitamin D is not associated with COVID-19, it still plays an important role in the maintenance of general health.

> *“@redacted the way I look at it, it doesn’t hurt to take them so why not. If it helps at all with covid, great. If it doesn’t, at least I’m getting my vitamin D.” (Twitter User 43)*
>
> *“@redacted Canadians should probably generally supplement with vitamin D in winter, unless you have regular blood work showing adequate levels (or some other medical reason). Little downside, potential big upside. Too bad a likely essential pre-hormone has become politicized by covid.” (Twitter User 44)*

### Ground Truth

Figure 2 shows the number of ‘correct’ and ‘incorrect’ codes identified when data from Twitter (12/2/2021 Tweets) were compared to the key findings from the NICE report titled “COVID-19 rapid guideline: vitamin D” (National Institute for Health and Care Excellence, 2020). The majority of the information relating to the key findings was ‘incorrect’ for all of the findings.

**Figure 2.**
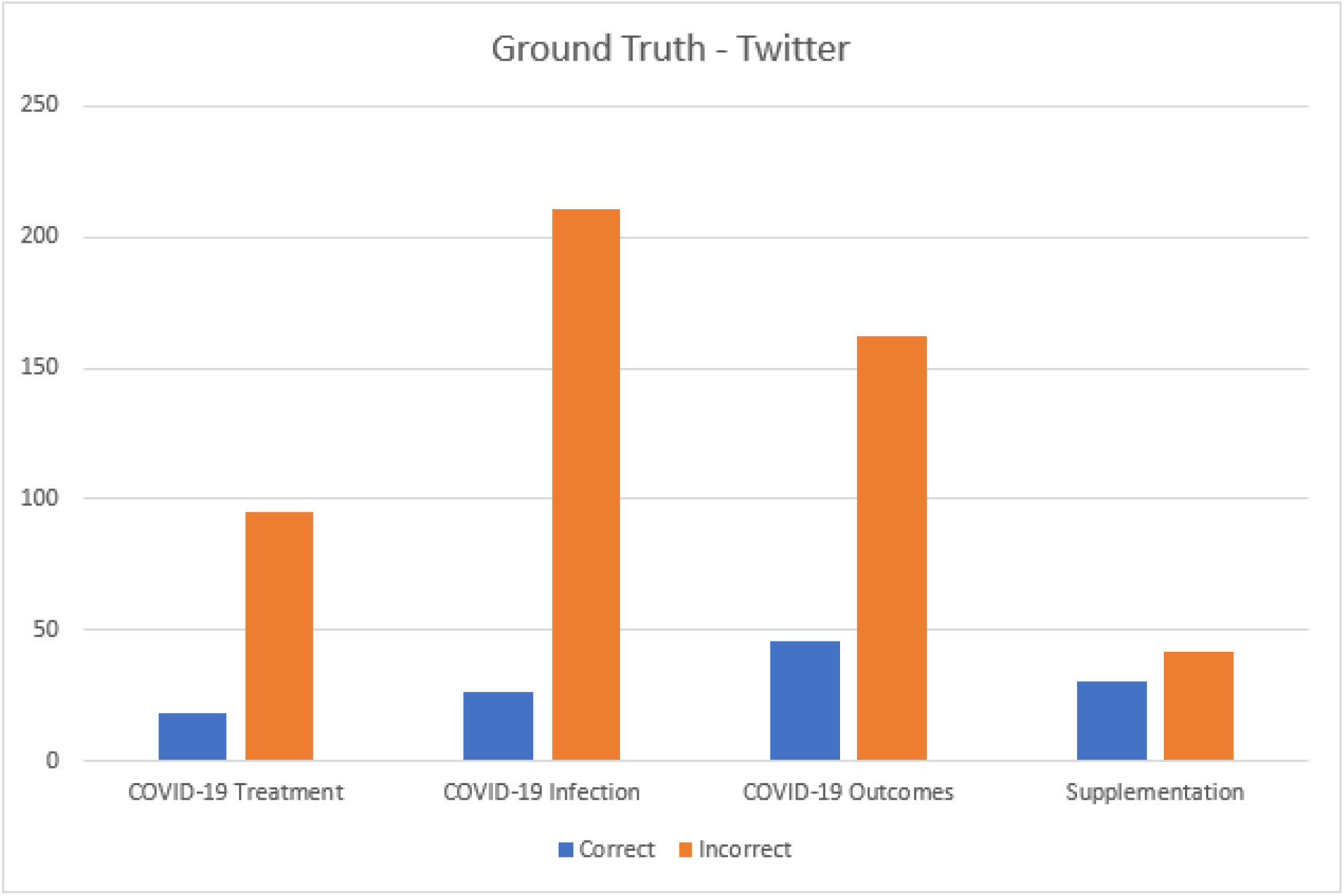
‘Correct’ vs ‘Incorrect’ opinions from Twitter.

### 1. COVID-19 Treatment

The NICE report states that vitamin D supplementation should not be used as a COVID-19 treatment. ‘Incorrect’ Tweets were over five times more common than ‘correct’ Tweets.

#### Correct

> *“@redacted There is no evidence that vitamin D has anything to do with COVID. It certainly doesn’t seem to have any impact on treating it.” (Twitter User 11)*

#### Incorrect

> *“I’m obviously thrilled with this study result, but I also feel pretty sad. It was pretty clear to me in August that vitamin D was a potent treatment for Covid, 1.5 million people have died since then.” (Twitter User 45)*

### 2. COVID-19 Infection

Vitamin D supplementation should not be offered to prevent COVID-19 infection. COVID-19 infection was the most discussed theme on Twitter. This theme also had the highest proportion of ‘incorrect’ information.

#### Correct

> *“@redacted They didn’t. They’ve been telling people to take vitamins for the last 50 years. Telling people to specially take vitamin D for COVID like it’ll make them immune when it won’t even come close is however a murderously bad idea in anyone with a brain’s book.” (Twitter User 46)*

#### Incorrect

“@redacted The HSE chiefs should have acted earlier and informed the public that vitamin D is a great preventative for covid. This needs to be addressed by the nurses union” (Twitter User 47)

### 3. COVID-19 Outcomes

Low vitamin D status is associated with more severe COVID-19 outcomes; however, it is not possible to confirm causality. ‘Incorrect’ Tweets were almost four times more common than ‘correct’ Tweets.

#### Correct

> *“The serum concentration of vitamin D falls during a systemic inflammation which may occur during severe covid-19 illness and it is difficult to know if low vitamin D status causes poor outcomes or vice versa.” (Twitter User 48)*

#### Incorrect

> *“@redacted I noticed they didn’t mention vitamin D deficiency in Black people. Which is the leading cause of mortality from Covid.” (Twitter User 49)*

### 4. Supplementation

Adults, young people, and children over 4 years should consider taking a daily supplement containing 10 micrograms of vitamin D between October and early March. Groups at high risk of deficiency should also consider year-round supplementation. Supplementation was the least discussed theme on Twitter. However, this theme had the highest proportion of ‘correct’ Tweets when compared to the other key findings.

#### Correct

> *“Surprised to discover that the NHS recommends *everyone* take Vitamin D from October to March, during Covid.” (Twitter User 50)*

#### Incorrect

> *“@redacted RDA is typically grossly underestimated (I wonder why that it is?). 5,000iu D3 to maintain your existing level. 10,000iu to increase your D3 level.” (Twitter User 39)*

## Discussion

This study identified how the relationship between vitamin D and COVID-19 has been represented on the social media site Twitter. The four main themes identified through inductive thematic analysis were association of “vitamin D with COVID-19”, “politically informed views”, “vitamin D deficiency” and “vitamin D sources” (Table 1). When compared to the ground truth, the majority of information relating to the key findings was ‘incorrect’ for all of the findings (Figure 2). The ground truth analysis carried out in this study is similar to the fact checking processes traditional journalists engage in as part of their professional practices, especially on social media (Juneström, 2020).

Considering the implications of these findings on public health, high levels of misinformation are dangerous in the context of COVID-19, as information is a key factor in disease prevention (Mejia et al., 2020). Ofcom reported that in 2020, 45% of people in the UK used social media to follow breaking news stories (Ofcom, 2021). Due to the large quantity of misinformation identified (Figure 2), it is likely that many in this portion of the population are exposed to similar misinformation on a regular basis. Consider one of the 45% reading the Tweet “@redacted The HSE chiefs should have acted earlier and informed the public that vitamin D is a great preventative for covid This needs to be addressed by the nurses union” (Twitter User 49). If they believe the Tweet contains correct information, they may stop following guidelines such as social distancing that aim to prevent COVID-19 infection, and instead begin supplementing with vitamin D. In this case, their risk of infection, and consequently the risk faced by others in the local community would increase. However, no data on behaviour change in the public was collected, so it is not possible to confidently draw any conclusions regarding the impact of the misinformation on public health. Though, similar previous research measuring the impact of COVID-19 vaccine misinformation on vaccination intent highlighted that exposure to online misinformation reduced intent to accept a vaccine (Loomba et al., 2021). Therefore, in this study, it is hypothesised that public health may have been negatively impacted by the high levels of misinformation shared on Twitter.

Comparing this study to previous research concerning social media response to infectious disease outbreaks, both the methodology and results are similar. (Ahmed et al., 2019) explored Twitter users views on the H1N1 virus by thematically analysing Tweets collected using keyword searches. Some of the 8 key themes and 38 subthemes identified by researchers are comparable to themes and subthemes identified in this study relating to vitamin D and COVID-19. Politics and food were two of the main themes, while prevention products, medication and media criticism were sub themes.

Another similar study, (Oyeyemi, Gabarron & Wynn, 2014) used keyword searches on Twitter to test the quality of Ebola related information shared by users in Guinea, Liberia, and Nigeria from 1st to 7th September 2014. It was reported that 55.5% of Tweets contained medical misinformation, while only 36% contained medically correct misinformation. Notably, the proportion of incorrect information relating to vitamin D and COVID-19 reported in this study is even more skewed (Figure 2). High levels of misinformation are synonymous with social media sites such as Twitter because users sharing ‘news’ do not need to adhere to the same verification processes as traditional journalists (Boididou et al., 2017). Consequently, users can freely share misinformation with no repercussions. Current Twitter policy states sharing content that may mislead people about “the efficacy and/or safety of preventative measures, treatments, or other precautions to mitigate or treat the disease (COVID-19)” may not be shared on Twitter (COVID-19 misleading information policy, 2021). However, it is evident that this policy is not being enforced effectively (Figure 2). Therefore, Twitter should consider improving their current methods of combating misinformation in order to effectively enforce their policy.

The strengths and weaknesses of this study must be considered. Collecting Tweets on a weekly basis provided a high volume of data and allowed researchers to track changes in the number of Tweets shared (Figure 1), providing a good insight into Twitter users behaviours throughout this time period. However, NCapture can only collect Tweets published in the previous eight days, meaning that Twitter data from the early stages of the pandemic were unavailable. It is possible that insights from Twitter would be different if data from earlier periods of the pandemic were available. Furthermore, data was only obtained from Twitter. Different social media platforms contain different types of information (Cinelli et al., 2020), so it is likely that if data were also analysed from other platforms that the results would differ. Additionally, as only textual data was collected, other types of data such as images, videos and emojis were not included in the analysis. Therefore, the nuance of other data types was not captured. It is also important to note that the only keywords used were “vitamin D” and “COVID”, meaning that Tweets using abbreviations such as “vit D” were not collected.

This study highlights the extent of the issue social media sites face with misinformation. Companies like Twitter have an obligation to fight misinformation, as it can be detrimental in disease prevention (Mejia et al., 2020). Future research should consider insights from various social media platforms. Different social media platforms contain different types of information (Cinelli et al., 2020), so it is possible that insights relating to COVID-19 may also differ.

## Data Availability

All data produced in the present study are available upon reasonable request to the authors

